# Inverse correlation between NLRP3 and TMEM176B as a possible tool for colorectal cancer progression

**DOI:** 10.1101/2024.08.30.24312793

**Authors:** Raylane Adrielle Gonçalves Cambui, Mariela Estefany Vera Roa, Vinicius Nunes Cordeiro Leal, Suemy Melim Yamada, Leonardo Antônio Teixeira de Oliveira, Izabel Nazira Nadaf, Cleiton Ribeiro Lelis, Rafaela Cássia da Cunha Pedroso, Rafaela Cardoso do Nascimento Costa, Raquel Maria Neves Amorim, Jefferson Rodrigues do Amaral, Polyana Silva Lemes, Leonardo Amorim Rizzo, Gilmar Ferreira do Espírito Santo, Rosa Maria Elias, Bruno Cogliati, Marcelo Hill, Alessandra Pontillo

## Abstract

**Purpose:** TMEM176B has been recently identified as a novel player in anti-cancer immune responses by negatively modulating the NLRP3 inflammasome activation in colorectal cancer (CRC). Yet, the TMEM176B/NLRP3 axis in CRC needs to be deeply investigated.

**Methods:** TMEM176B and NLRP3 expression were evaluated in CRC patients by immunohistochemistry and RT-qPCR assays. The prognostic relevance of TMEM176B and NLRP3 was determined by in silico analysis. The NLRP3 inflammasome activation in the CRC microenvironment was assessed in a peripheral blood mononuclear cells (PBMC) and CRC cell line (HCT-116) spheroids co-culture assay.

**Results:** Reduced NLRP3 and increased TMEM176B expression correlates to CRC stages and poor survival. The in vitro assay showed HCT-116 cells activated NLRP3 inflammasome in PBMC.

**Conclusion:** These findings evidence the inverse correlation between NLRP3 and TMEM176B in CRC progression, suggesting them as a predictive tool.

## Introduction

The NLRP3 inflammasome apparently displays a dual and ambiguous role in colorectal cancer (CRC): on one hand, it can foster an inflammatory microenvironment that promotes tumor growth, and on the other, it can activate immune responses that target and eliminate cancer cells (Deng et al., 2023). Several studies based on the Azoxymethane (AOM)/Dextran Sodium Sulfate (DSS) mouse model of inflammatory CRC have proposed a protective function of the NLRP3 inflammasome in preventing colitis and intestinal tumorigenesis (Zaki et al., 2010; Allen et al., 2010; Dupaul-Chicoine et al., 2015; Segovia et al., 2019). In contrast, different investigations have characterized NLRP3 as a potential risk factor for the development of both conditions (Bauer et al., 2010; Huber et al., 2012). In humans, CRC patients with lower levels of NLRP3 expression showed a more favorable prognosis (Shi et al., 2021; Wang et al., 2020).

According to these contrasting findings, in recent genetic association studies we demonstrated that a gain-of-function (GoF) missense variant in the NLRP3 (Asp703Lys) and a loss-of-function (LoF) missense variant in its negative regulator TMEM176B (Ala134Thr) were associated to unfavorable or favorable prognosis, respectively (Cambui et al., 2020; Cambui et al., 2023).

The NLRP3 inflammasome, serves as a sensor for cellular stress and damage, triggering the release of pro-inflammatory cytokines such as interleukin (IL)-1β and IL-18, as well as the inflammatory type of cell death called pyroptosis (Barnett et al., 2023 and Wang et al., 2023). The NLRP3 receptor has been detected in several cell types within the intestinal mucosa, including its expression in epithelial cells and resident macrophages (Song-Zhao et al., 2014; Kummer et al., 2007; Seo et al., 2015), where it plays a crucial role for maintaining intestinal balance. The activation of NLRP3 inflammasome in the myeloid lineages can positively impact the adaptive immune response by triggering anti-tumoral CD8+ T cells (Segovia et al., 2019). On the other hand, an imbalance between IL-1ß and IL-18 may impact the CD4+ T cells polarization (Th17 versus Th1/Th2) in tumor microenvironment and affect the anti-cancer immune response (Van Den Eeckhout et al., 2015). Otherwise, the NLRP3 inflammasome is also expressed by non-immune cells, such as epithelial and endothelial cells and fibroblast, that compose the tumor microenvironment and can also play divergent roles in tumor growth (Yang et al., 2019).

Among the proteins influencing the NLRP3 inflammasome activation (Yamada and Pontillo, 2022), TMEM176B stands out since it was recently discovered as a negative regulator of the NLRP3. TMEM176B is an ubiquitous non-specific cation channel highly expressed in primary and secondary lymphoid organs, colon, lung and liver (Hill et al., 2022). Both knockout and pharmacological inhibition of TMEM176B reduce tumor growth and enhance survival rates in animals inoculated with tumoral cell lines MC38 (colon), LL/2 (LLC1; lung), or EG7 (thymic lymphoma) (Segovia et al., 2019), suggesting that, at least in these models, a reduced TMEM176B expression/function - and the consequent increased activation of the downstream NLRP3 pathway - associates with increased survival. Since TMEM176B is a draggable molecule, it represents a possible target for therapies aimed to increase NLRP3 inflammasome-mediated anti-tumor response.

Here, we explore the expression of NLRP3 and TMEM176B in the tumoral tissue of CRC patients, to deeper characterize their correlation taking in account the cell-type specific expression (tumor cells, leukocytes aggregates) and the patient clinical stage (early: I-II, late: III-IV).

## Patients and Methods

### Patients and healthy donors

Fifteen CRC patients were recruited at the “Instituto de Tumores e Cuidados Paliativos de Cuiabá do Hospital Geral e Maternidade de Cuiabá” and the “Clínica de Tratamento Multidisciplinar do Câncer” (ONCOMED) in Cuiabá. (MT, Brazil). CRC was diagnosed based on histopathologic characteristics and staged according to the TNM system (TNM I to IV) as endorsed by the Union for International Cancer Control (UICC) (2010) (**Table 1**). Six healthy donors (HD) who are matching for sex and age with the patients’ group (median: 57 years; min-max: 43-74) were included as a control group for some experiments. The Ethics Committee of ICB/USP (CEPSH) (process number 30173919.1.0000.5467) approved the study.

**Table 1.**
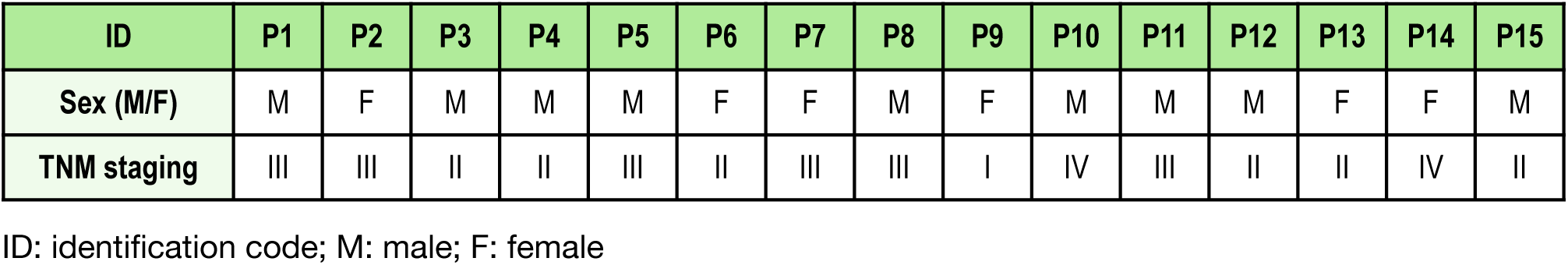
Patients’ Characteristics.

| ID | P1 | P2 | P3 | P4 | P5 | P6 | P7 | P8 | P9 | P10 | P11 | P12 | P13 | P14 | P15 |
| --- | --- | --- | --- | --- | --- | --- | --- | --- | --- | --- | --- | --- | --- | --- | --- |
| Sex (M/F) | M | F | M | M | M | F | F | M | F | M | M | M | F | F | M |
| TNM staging | III | III | II | II | III | II | III | III | I | IV | III | II | II | IV | II |

ID: identification code; M: male; F: female

### Immunohistochemistry analysis

Formalin-fixed paraffin-embedded (FFPE) tissue blocks from the most prominent tumor areas of twelve CRC patients (P1-P12) were stained for NLRP3 and TMEM176B using the NovoLink kit (Leica Biosystems). Positive reactions were revealed by DAB staining (DAB Chromogen, NovoLink kit). NLRP3 and TMEM176B staining were assessed through a semi-quantitative method (Ruifrok e Johnston; 2001) using the “Colour Deconvolution 2” plugin of ImageJ (Landini et al., 2021). The histological HE analysis in early-stage cases exhibited well-defined glandular structures with uniform nuclei and minimal cytological atypia (**Supplementary File 1A**). In contrast, the advanced-stage cases revealed disorganized glandular architecture, indicating tumor invasion into the adjacent layers of the tissue (**Suppl.File 1B)**. Additionally, features such as an increased nucleus-to-cytoplasm ratio, nuclear atypia, and frequent mitoses were observed, indicative of high proliferative activity. Microscopic analysis also showed the presence of an inflammatory infiltrate at the tumor site (**Suppl.File 1B**). The architecture of the glands in normal mucosa appeared relatively preserved, indicating limited invasion into the surrounding tissue layers (**Suppl.File 1C**). The nucleus-to-cytoplasm ratio was within normal limits, and mitotic figures were infrequent. Moreover, infiltration of inflammatory cells was minimal.

### RNA isolation and expression analysis

Total RNA was isolated from fresh tumor biopsies (about 0.2 cm in size) and peripheral blood (4 mL) of six CRC patients (P1-P3, P13-P15) and healthy donors (n = 6), as well as from HCT-116 cells using the RNeasy Mini Kit (Qiagen), following the manufacturer’s instructions. After reverse transcription (SuperScript™ III Reverse Transcriptase ki; Invitrogen, ThermoFisher Scientific), the cDNA was used for gene expression analysis by qPCR and gene-specific Taqman® assays (Thermo Fisher Scientific) **(Supplementary File 2A)** utilizing the QuantStudioR 3 real-time (RT) PCR system (Thermo Fisher Scientific). The QuantStudio 3.0 software was utilized to obtain cycle threshold values (Ct). Basal (constitutive) gene expression was normalized by the Ct value of housekeeping gene *GAPDH* and then reported as 2^(-ΔCt) (Schmittgen and Livak, 2008).

### In vitro assays

The human colorectal cancer cell line HCT-116 (ATCC® CCL-247™) (kindly donated by Prof. Leticia Lotufo, Laboratory of Marine Pharmacology, ICB/USP) was used for spheroids formation. 1.8×10^5^ HCT-116 cells were seeded in U-shaped 96-well plates (Nuclon Sphera, Thermo Fisher) and incubated for 72 hours (Leek, 2016; Stone, 2019). Peripheral blood mononuclear cells (PBMC) were isolated from fresh blood of three HD by using Ficoll solution (GE Healthcare, Merck) and added to HCT-116 spheroids (1:1) in a fresh culture medium for 24 hours. Cells were harvested and incubated with the PerCP-Cy5.5 anti-CD45 antibody and the FAM-FLICA® Caspase-1 (YVAD) Assay Kit (Immunochemistry technologies) following the manufacturer’s instructions. Cells were then incubated with LIVE/DEAD Dead Cells Stain (Thermo Fisher Scientific) and analyzed by flow cytometry (BD Canto II; BD Biosciences) (Gate strategy in **Supplementary File 3**). IL-1ß, IL-18 and LDH release was measured in culture supernatants by commercial kits (R&D Systems, Biolegend; Thermo Fisher Scientific). In some experiments cells were alternatively pre-treated with the NLRP3 inhibitor MCC-950 (10 µM, Invivogen) for 30 minutes or the TMEM176B inhibitor Bay-K8644 (Sigma-Aldrich) was added in LPS-primed cells for 15 minutes.

### Bioinformatics analysis

The prognostic relevance of TMEM176B and NLRP3 expression in CRC was assessed using the Kaplan–Meier Plotter database (http://kmplot.com/analysis/) (Gyorffy, 2024). The correlation between TMEM176B and NLRP3 mRNA levels and overall survival (OS) in the CRC group was calculated by the Kaplan–Meier curve and log-rank test, a tool that automatically generates risk ratios (HR) and 95% confidence intervals, with a p value of < 0.05 considered statistically significant.

### Statistical Analysis

Statistical analyses in our study were conducted using GraphPad Prism 8.0. Quantitative data were evaluated using two-tailed Student’s t-test or one-way ANOVA. Data are expressed as the mean and standard deviation (SD) of at least three independent experiments. Statistical significance was determined at *p < 0.05, **p < 0.01, ***p < 0.001. Non-significant results are not indicated in the graphs.

## Results

In our previous association studies (Cambui et al., 2020 and 2023) we have shown that common variants in *NLRP3* and its regulator *TMEM176B* play a role in the individual susceptibility to CRC prognosis. However the cells and tissues mainly affected by genetic predisposition and the effect on CRC prognosis are not clearly elucidated. Therefore here we dissected the TMEM176B/ NLRP3 axis by the use of different approaches.

First we analyzed the expression of these two proteins in paraffined samples from twelve samples of CRC at different stages (TNM I-IV). All the studied samples (n = 12; P1-P12) resulted positive for NLRP3 staining, however we noted that the local NLRP3 expression is higher in advanced stages (TNM III/IV: P1, P2, P5, P7, P8, P10, P11) compared to early stages (TNM I/II) (P3, P4, P6, P9, P12) (**Figure 1 A-B**). When we compared the staining signal for each patient and groups we observed that the difference between TNM I/II and III/IV in NLRP3 expression in the total area is statistically significant (p = 0.038) **(Fig.1 C**).

**Figure 1.**
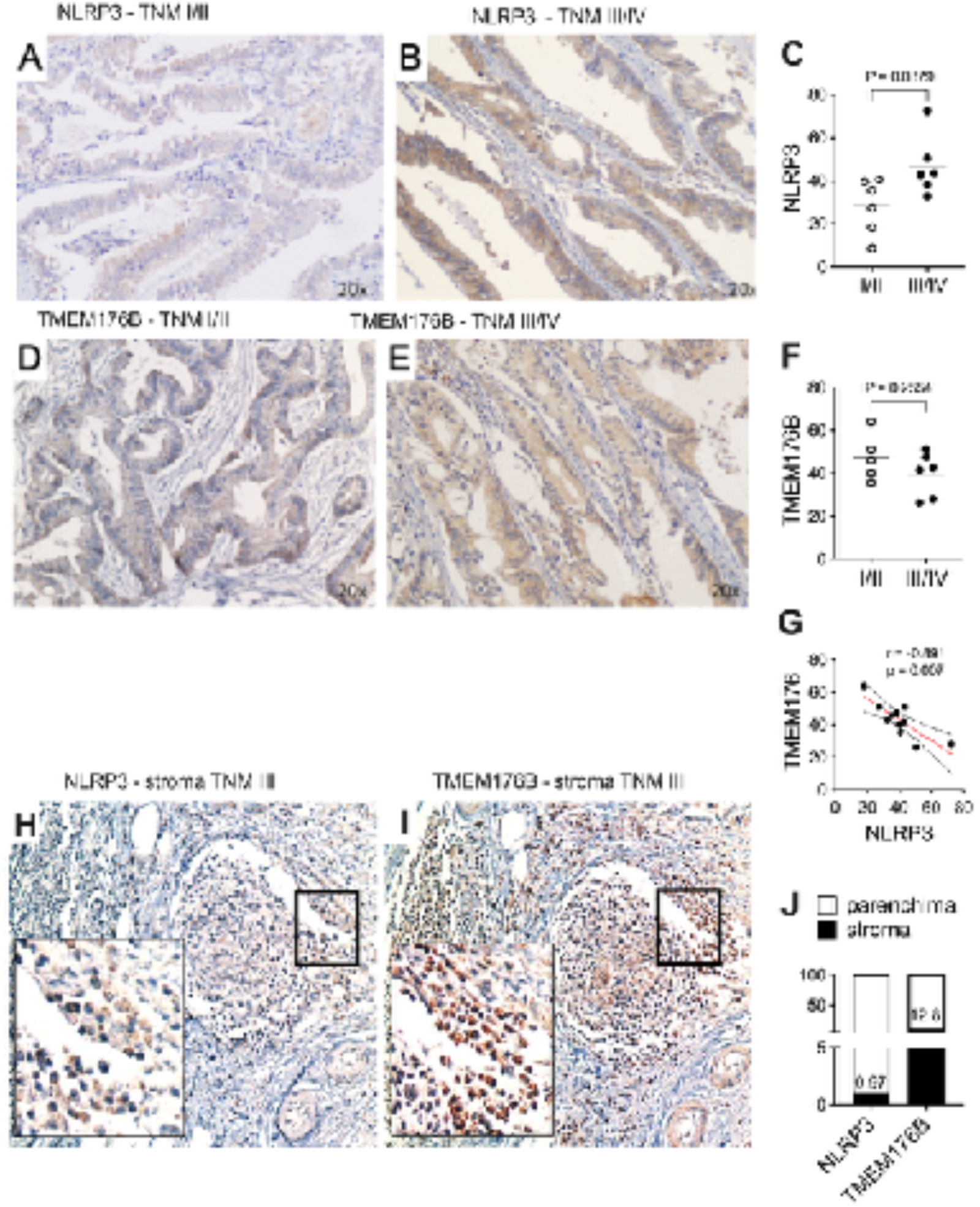
Inverse expression of TMEM176B and NLRP3 in CRC biopsies. (A-B) NLRP3 staining in formalin/PFA-fixed paraffin-embedded CRC tissue in one early-stage (TNM I) and advanced-stage (TNM III). NLRP3 is stained with 3,3’-diaminobenzidine tetrahydrochloride (DAB) chromogen (brown) and cell nuclei stained with hematoxylin (blue). The images were captured at 20x magnification. (C) Scatter plot shows the percentage of NLRP3 staining for all the CRC biopsies analyzed (n =12) grouped according to the stages (TNM I/II versus TNM III/IV). Unpaired two-tails t-test p-value is reported. (D-E) TMEM176B staining in formalin/PFA-fixed paraffin-embedded CRC tissue in one early-stage (TNM I) and advanced-stage (TNM III). NLRP3 is stained with 3,3’-diaminobenzidine tetrahydrochloride (DAB) chromogen (brown) and cell nuclei stained with hematoxylin (blue). The images were captured at 20x magnification. (F) Scatter plot shows the percentage of TMEM176B staining for all the CRC biopsies analyzed (n =12) grouped according to the stages (TNM I/II versus TNM III/IV).Unpaired two-tails t-test p-value is reported. (G) Correlation analysis for NLRP3 and TMEM176B staining in CRC biopsies. Pearson correlation coefficient (r) and p-value (p) are indicated. (H-I) NLRP3 and TMEM176B staining in formalin/PFA-fixed paraffin-embedded CRC tissue in one advanced-stage (TNM III) with focus on stromal region. NLRP3 or TMEM176B are stained with 3,3’-diaminobenzidine tetrahydrochloride (DAB) chromogen (brown) and cell nuclei stained with hematoxylin (blue). The images were captured at 20x and 100x magnification (minor rectangle in the left bottom part of the images). (J) Stacked bar graph shows NLRP3 or TMEM176B percentage of staining in the stroma. The percentage value of cells positive for staining is reported.

On the other side, TMEM176B exhibited mild expression in CRC tumors (**Fig. 1D-E**), and without significant difference between TNM I/II and TNM III/IV groups (p = 0.255) (**Fig. 1F**). TMEM176B is a negative regulator of NLRP3, therefore we examined whether a correlation exists between the expression of the two molecules. TMEM176B and NLRP3 exhibit a negative correlation **(Fig. 1G)**, which is maintained only in TNM I/II group, when we separately analyzed the TNM I/II and III/IV groups (data not shown).

Segovia and colleagues (Segovia et al., 2019) have reported that there is a differential expression of TMEM176B in tumor biopsies when taking in account the stroma. Thus, a high stromal TMEM176B expression in CRC was associated with reduced patient survival. They have suggested that the increased expression of TMEM176B affects the NLRP3 level in immune cells infiltrating the tumor, such as macrophages. To assess this condition, we analyzed the expression of TMEM176B and NLRP3 in our TNM III CRC samples (n = 6) focusing on the leukocytes. As expected, we observed an enriched leukocyte area in the sample with a weak staining for NLRP3 (**Fig.1 H, J**) and a mild-to-strong signal for TMEM176B (**Fig.1 I-J)**. Therefore, our findings are in accord to what previously observed (Segovia et al., 2019) and add a novel concept about the NLRP3 expression in CRC. Since it is supposed that leukocytes constitutively expressed the NLRP3 receptor, these findings suggest that as the NLRP3 is highly expressed in TNM III tumors compared to lower TNM stages, this positiveness is not related with leukocytes infiltration, suggesting that at advanced CRC stages the NLRP3 may be mainly expressed in other cell types.

Then we wondered whether the protein expression measured by immunohistochemistry could be predicted by RNA expression level in the tumor biopsy and/or peripheral blood, as the most accessible sample in routine exams. Therefore, we obtained fresh biopsies and whole blood of six CRC patients (P1-3, P13-15) and evaluated the NLRP3 and TMEM176B gene expression by qPCR. For the blood assay we also included a group of healthy donors (HD; n = 3). In tumor biopsies the expression level of NLRP3 is low (**Figure 2A**) and is not significantly different between TNM I/II and TNM III/IV groups (**Supplementary File 4A**). Even if it is expressed at a higher level than NLRP3, TMEM176B is not differently expressed between the two groups (**Fig. 2A; Suppl. File 4B**). In whole blood, the expression level of NLRP3 and TMEM176B is comparable, without significant differences between TNM groups or with HD (**Fig. 2B; Suppl. File 4 D-H)**. The expression level of other components of the inflammasome also did not differ between the TNM groups (**Suppl. File 4 C, 1F**). Of note, the plasma level of the cytokine IL-18 (not IL-1ß) was higher in CRC patients than in HD (p = 0.0006) (**Fig. 2C-D**), suggesting that even if we cannot observe a different expression profile of inflammasome genes, the inflammasome is more activated in patients compared to HD.

**Figure 2.**
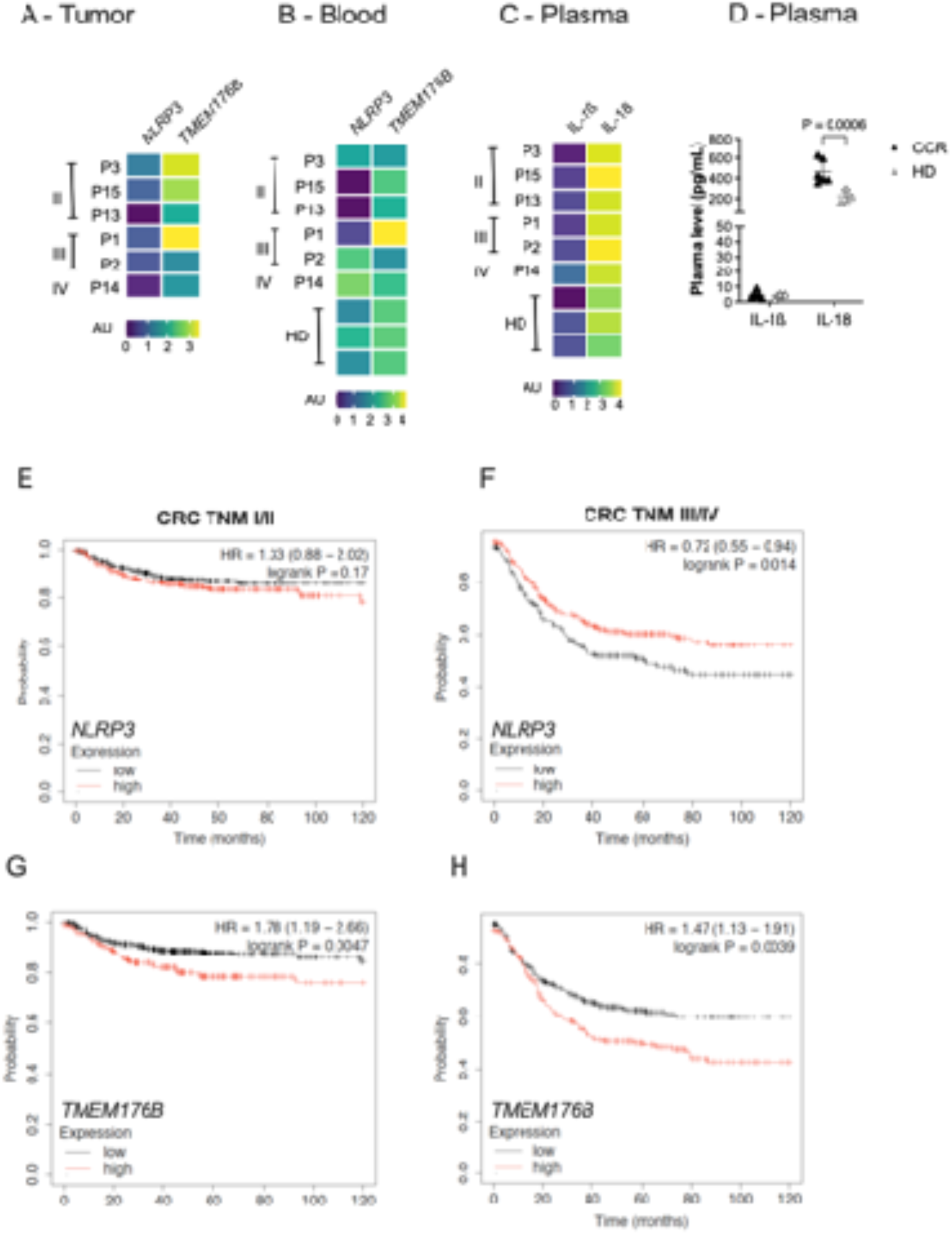
Transcriptional level of NLRP3 and TMEM176B varied according to CRC stage and prognosis. A-B) Heatmap graphs show RNA expression level of NLRP3 and TMEM176B genes in fresh tumor biopsies and peripheral blood leukocytes of six CRC patients, three with TNM-II, two with TNM-III and one with TNM-IV. Values are reported as log (2exp-ΔCt). C-D) Heatmap and scatter plot graphs show plasma levels of IL-1ß and IL-18 of six CRC patients, three with TNM-II, two with TNM-III and one with TNM-IV. In heatmap values are reported as log (pg/mL). Unpaired two-tails t-test p-value is reported. (D-E) Kaplan–Meier curves for CRC patients survival according to NLRP3 expression level in TNM-I/II and TNM-III/IV stages. (F-G) Kaplan–Meier curves for CRC patients survival according to TMEM176B expression level in TNM-I/II and TNM-III/IV stages. Data and curves were obtained from the Kaplan–Meier Plotter database (http://kmplot.com/analysis/). Hazard Ratio (HR) and long rank P value are reported.

Even if we were not able to identify significant differences in genes expression according to the CRC staging, in a public database we can observe that increased NLRP3 expression was associated with increased survival in TNM-III/IV CRC (**Fig. 2E-F**). A reduced TMEM176B expression seems to be beneficial for the patient, independently from the tumoral stage (**Fig. 2G-H**).

Finally, we developed a 3D cell culture model (cancer spheroids) to imitate a tumoral microenvironment to deeper investigate the role of NLRP3 and TMEM176B in cancer cells and leukocytes.

We analyzed the NLRP3 inflammasome activation in co-cultures of colorectal cancer cell line HCT-116 spheroids and peripheral blood mononuclear cells (PBMC), trying to depict if the complex could be triggered in the tumor itself and/or in immune infiltrate. The co-culture itself can increase caspase-1 activation (**Figure 3 A-B**) as well as the release of IL-1β (but not of IL-18) and LDH, which are not detected in PBMC or spheroids alone (**Fig.3 B-D**).This heightened activation of the inflammasome in the co-culture condition was significantly reduced when cells were treated with the specific NLRP3 inhibitor MCC-950 (**Fig.3 B-D**), indicating that the co-culture induces specifically the NLRP3 inflammasome activation.

**Figure 3.**
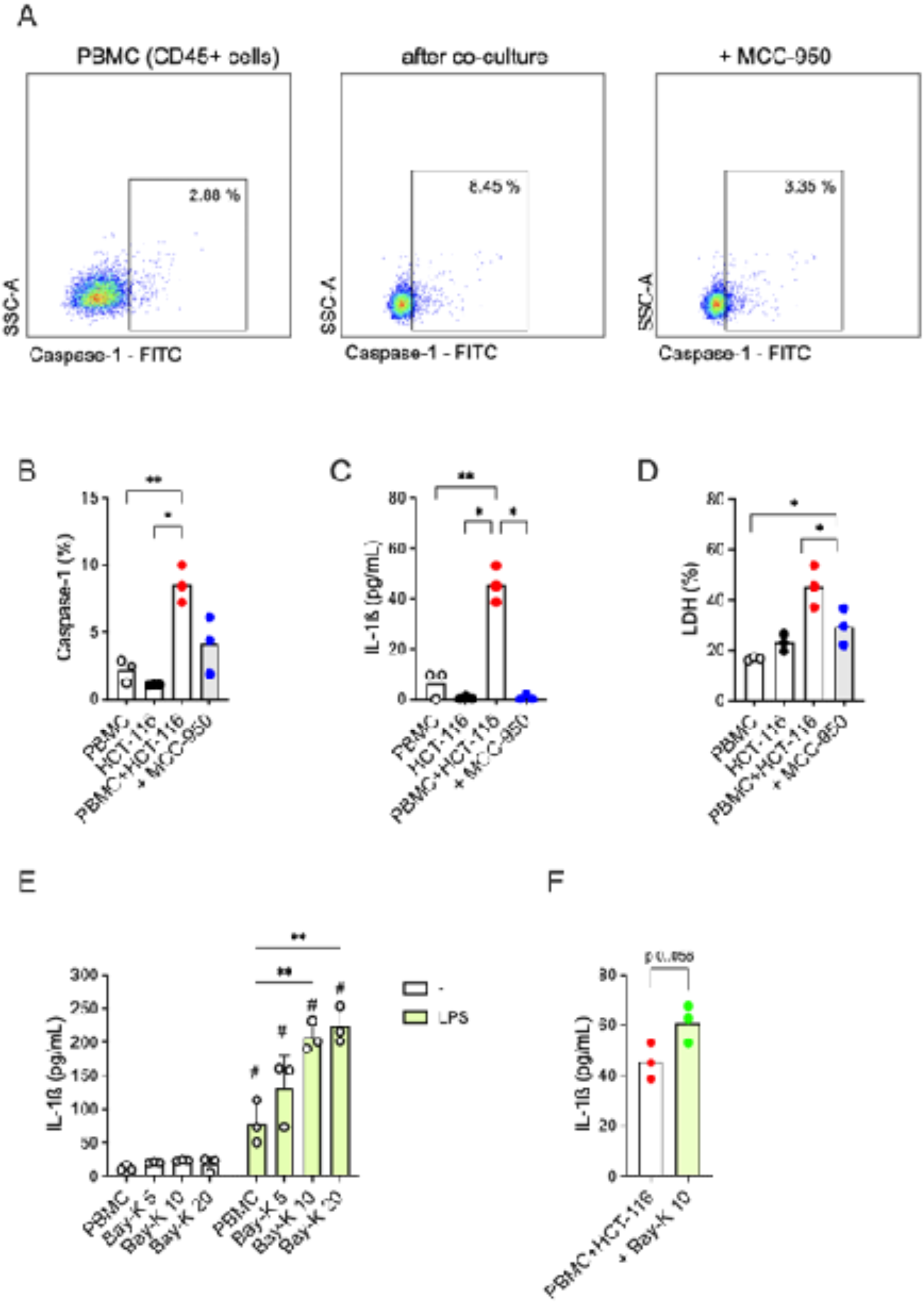
Co-culture of HCT-116 spheroids with PBMC induced NLRP3 inflammasome activation in PBMC. HCT-116 spheroids (1.8 x10^5) were cultured alone or with PBMC (1.8 x10^5 cells, HCT-116/PBMC ratio: 1/1) isolated from three healthy donors for 24 hours, eventually with the pre-treatment of the NLRP3 inhibitor MCC-950 (10 µM) for 30 minutes. PBMC were harvested for activated caspase-1 detection using FAM-FLICA kit and flow cytometry. IL-1ß and LDH release was measured in culture supernatants. **A)** A representative experiment (out of three) of flow cytometry analysis of activated caspase-1 by the use of FAM-FLICA kit. Percentage of caspase-1 positive cells is reported for PBMC alone, PBMC from co-culture assay with HCT-116 and with the pretreatment with MCC-950 (10 µM). Gate strategy is reported in **Supplementary File 3**. **B-D)** Scatter dot plots with bars (mean) reports (B) the percentage of PBMC positive for activated caspase-1, **(C)** the concentration of IL-1ß and (**D)** the percentage of LDH released. **(E)** PBMC (1.8 x10^5 cells) isolated from three healthy donors were treated or not with LPS 1 µg/mL for 24 hours and then with the TMEM176B inhibitor BayK-8664 at increasing concentrations (5, 10, 20 µM) for 15 minutes. IL-1ß release was measured in culture supernatants. **(F)** HCT-116 spheroids (1.8 x10^5) were cultured with PBMC isolated from three healthy donors for 24 hours, with or without the treatment with Bay-K8644 (10 µM) for 15 minutes. IL-1ß release was measured in culture supernatants. **(B-D)** Paired One-Way ANOVA followed by multi comparison post-test was used to compare the conditions. *: p < 0.05; **: p < 0.01. **(E)** Paired Two-Way ANOVA followed by multi comparison post-test was used to compare the conditions within and between the two groups. **: p < 0.01 (within the group); #: p < 0.01 (between the groups). **(F)** Paired t test was used to compare the conditions.

It is interesting to emphasize that HCT-116 cells express a very low amount of *NLRP3, CASP1* and *IL1B* genes, even after LPS priming (**Suppl. File 2B**), therefore supporting the hypothesis that the immune cells, and not the spheroids, are responsible for inflammasome activation in the co-culture assay.

In mice, the TMEM176B inhibitor Bay-K8664 triggers caspase-1 activation and IL-1ß release in LPS-treated bone marrow-derived dendritic cells, which has been correlated with an increased CD8+T cells antitumoral activity and in turn may explain the effect of Bay-K8644 on tumor growth *in vivo* (Segovia et al., 2019). In our previous study, we reported a significant effect of loss-of-function variant *TMEM176B* rs2072443 on IL-1ß release in primary human monocytes, monocyte-derived macrophages and, partially, in lymphocytes (Cambui et al., 2023). Taking in account that all these cells express components of the NLRP3 inflammasome, these findings suggest a role of TMEM176B on NLRP3 modulation in leukocytes other than DC. To assess whether the TMEM176B could be involved in the NLRP3 inflammasome regulation in our *in vitro* model, we first treated the PBMC with increasing concentration of Bay-K8664 alone or in LPS-treated cells. As expected Bay-K8664 was not sufficient to induce alone any IL-1ß release in PBMC (**Fig. 3E**). However, concentrations up to 10 µM induced cytokine release in LPS-treated PBMC (**Fig. 3E**).

Then we conduct the HCT-116 and PBMC co-culture assay pre-treating PBMC with Bay-K8664 10 µM and observed an increase in IL-1ß release (**Fig.3 F**), compatible with the inhibitory effect on TMEM176B and activating effect on NLRP3 inflammasome.

## DISCUSSION

Several studies across various cancer types have consistently shown that increased infiltration of immune cells correlates with a more favorable clinical outcome, including in colorectal cancer (Galon et al., 2006-2007). Despite the acknowledged impact of the inflammasome on the recruitment and activity of immune cells, only a limited number of studies have explored its role in CRC prognosis. Some studies have highlighted the negative impact of NLRP3 on CRC (Shi et al., 2021; Wang et al., 2020). Accordingly, we previously showed that genetic variations in the NLRP3 gene, which lead to increased inflammasome activation, are associated with an unfavorable prognosis (Cambui et al., 2020). In line with this negative role of NLRP3, a loss-of-function variant (Ala134Thr) in TMEM176B has been found to be a protective factor for a favorable prognosis (Cambui et al., 2023). However, up to now, there is no data available that can explain these controversial findings and support the genetic association study.

Here we demonstrated that CRC biopsies exhibited varying expression of NLRP3 depending on the TNM staging, with lower levels in early-stage disease (TNM I/II) compared to advanced stages (TNM III/IV). Although several studies have shown that high NLRP3 expression is associated with worse prognosis in CRC (Shi et al., 2021; Wang et al., 2020) and other cancer types, such as gastric cancer (Wang et al., 2022), breast cancer (Saponaro et al., 2021), and pancreatic cancer (Zheng and Liu, 2022), our in silico analysis showed a significant beneficial effect of high NLRP3 expression on CRC patients survival, especially in the TNM III/IV group. These results suggest a potential protective role of NLRP3 expression in these stages of CRC. It is worth mentioning that the favorable influence of NLRP3 expression on cancer prognosis has also been reported for skin cutaneous melanoma (Wu et al., 2021).

In line with previous findings (Segovia et al., 2019), TMEM176B displayed uniform expression across all CRC cases, independently of the TNM classification, and its high level is significantly associated with poor survival, again independently of the TNM classification. The expression levels of NLRP3 and TMEM176B were significantly inversely correlated and significantly correlated with patients survival, suggesting the possibility of using these markers as a promising prognostic tool.

Given that biopsies are invasive and usually collected during surgery or, depending on the country’s health system, in CRC screening programs, we proposed analyzing blood samples for NLRP3 and TMEM176B expression to find a possible correlation with CRC. However, we could not detect a significant correlation between blood and CRC biopsies in terms of NLRP3 and TMEM176B expression. On the other hand, it is important to stress that plasma levels of IL-18 are increased in these patients, and even if this is not a specific marker, it could be considered, together with other signs, as a pathological fact.

Segovia et al. (2019) have also reported high stromal TMEM176B expression in CRC, suggesting that it can negatively regulate NLRP3 inflammasome activation. Accordingly, a detailed examination of the tumor-associated leukocyte aggregates (in our samples, we were able to detect them only in the advanced stages) revealed robust expression of TMEM176B, whereas NLRP3 was markedly scarce in this cellular compartment. Even if the expression levels of the two proteins do not indicate a functional cause-effect relationship, these findings lead us to hypothesize that in leukocytes, which commonly express TMEM176B (Hill et al., 2022), NLRP3 is downregulated. Indeed, leukocytes are also expected to express NLRP3; however, in our samples, they show low levels of NLRP3 at least in advanced stages of CRC. On the other hand, as we showed that advanced stages express significantly higher levels of NLRP3 than early ones, we wonder in which cell type this receptor is induced during CRC progression.

In our in vitro model of HCT-116 and PBMC, we observed that the co-culture itself (resembling the tumoral microenvironment) can induce NLRP3 activation in PBMC. We cannot relate this result to the histochemical data in terms of TNM stage, as we cannot determine whether we are mimicking a TNM I/II or III/IV microenvironment. However, we can hypothesize that at least in the early stages, the NLRP3 inflammasome can be activated by the tumor, and this response might help control its growth. During progression, possibly TMEM176B is up-regulated in leukocytes, negatively affecting the NLRP3-mediated response. Deeper investigations are needed to define the cause-effect relationship between these two molecules.

## Data Availability

All data produced in the present study are available upon reasonable request to the authors

## Acknowledgment

We thank all patients and their families and all personnel involved in patients’ attendance; Prof. Ana Paula Lepique Department of Immunology, Institute of Biomedical Sciences, University of Sao Paulo) for the help with spheroids and critical discussions, Prof. Anderson de Sa Nunes (Department of Immunology, Institute of Biomedical Sciences, University of Sao Paulo) for the use of ELISA Reader, Prof. Leticia Lotufo (Institute of Biomedical Sciences, University of Sao Paulo) for HCT-116 cell line.

## Disclosure Statement

No potential conflict of interest was reported by the authors

## Fundings

This study was supported by the Sao Paulo State Research Foundation (FAPESP) grant number 2019/06363-4 (A.P.) and 2021/0540-4 (A.P.). Authors are supported by fellowship programs: Excellence in research Fellowship from National Council for Scientific and Technological Development (CNPq) (A.P.); Post-doctoral fellowship from FAPESP (process n. 2020/15323-3) (V.N.C.L.); CAPES PhD Fellowship (R.A.G.C.; M.E.G.V.R.).

---

**Supplementary File 1.**
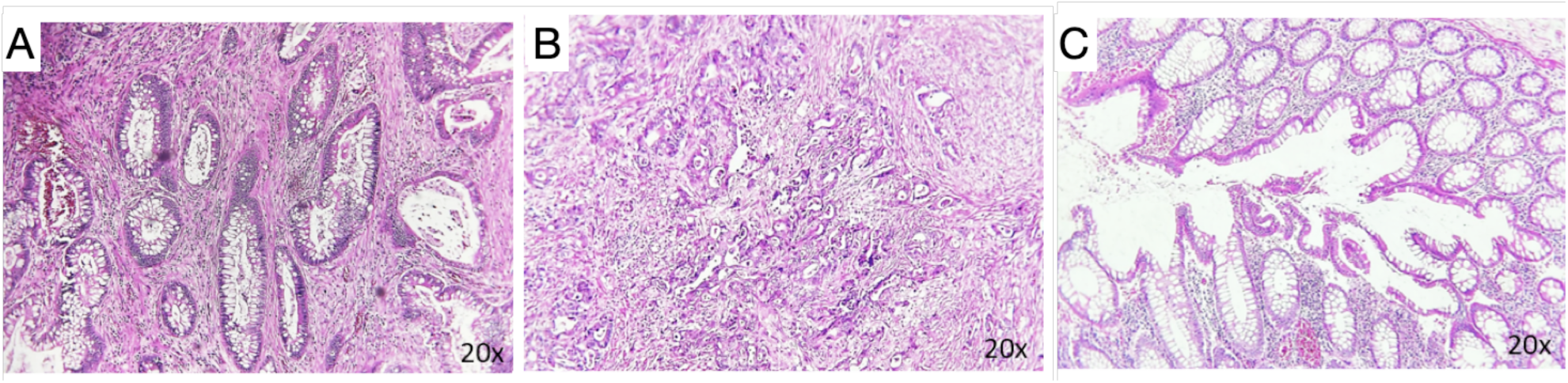
Hematoxylin and Eosin (HE) staining of CRC biopsies. A-B) Hematoxylin and Eosin (HE) staining in one early-stage (TNM I) and in one advanced-stage CRC (TNM III) cases. C) HE staining in normal adjacent intestinal epithelial tissue. The images were captured at 20x magnification.

**Supplementary File 2.**
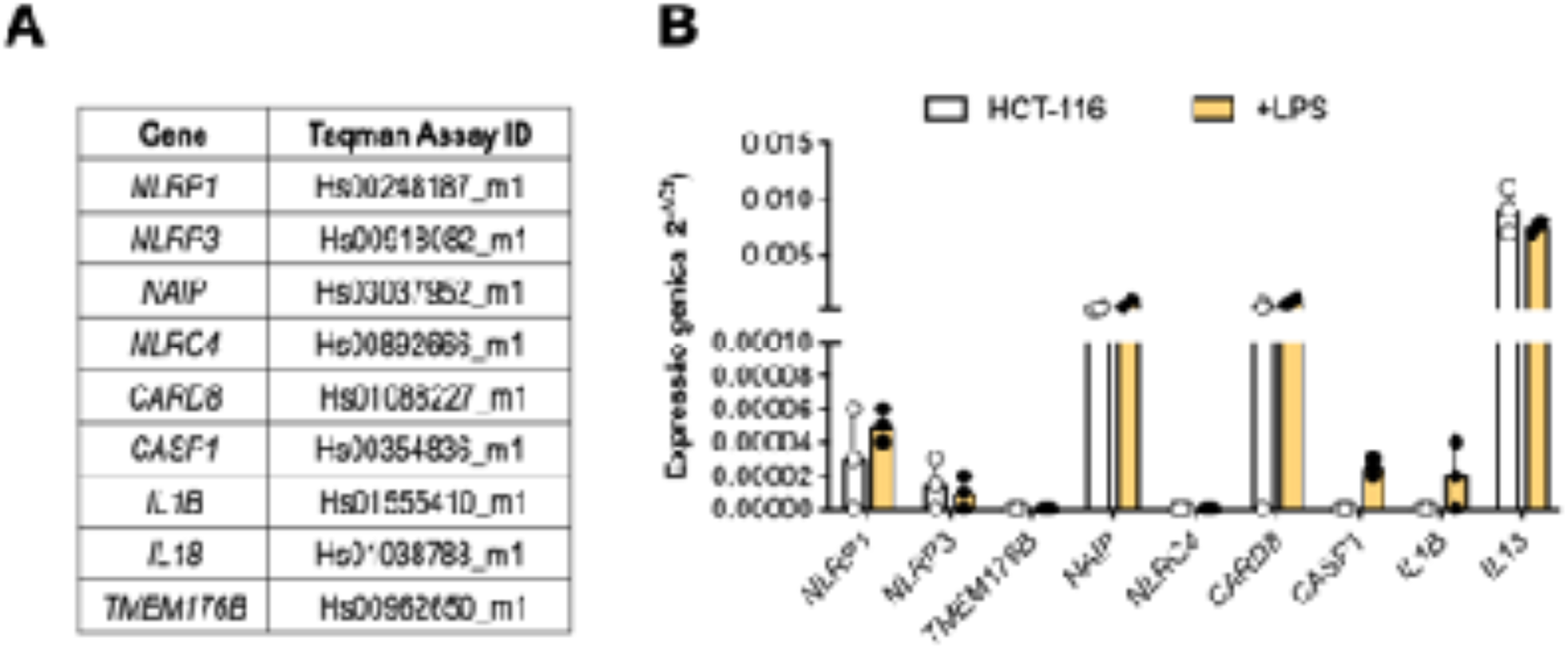
Relative Expression of inflammasome genes in HCT-116 cells. A) List of Taqman Gene expression assays for selected inflammasome genes. B) Relative gene expression of inflammasome genes in HCT-116 cell (0,125x10 exp6 cells) untreated and treated with LPS 1 µg/mL for four hours. The relative expression is reported as 2 exp -ΔCt, where ΔCt is the expression of target gene normalized for endogenous gene GAPDH. Three independent experiments were reported.

**Supplementary File 3.**
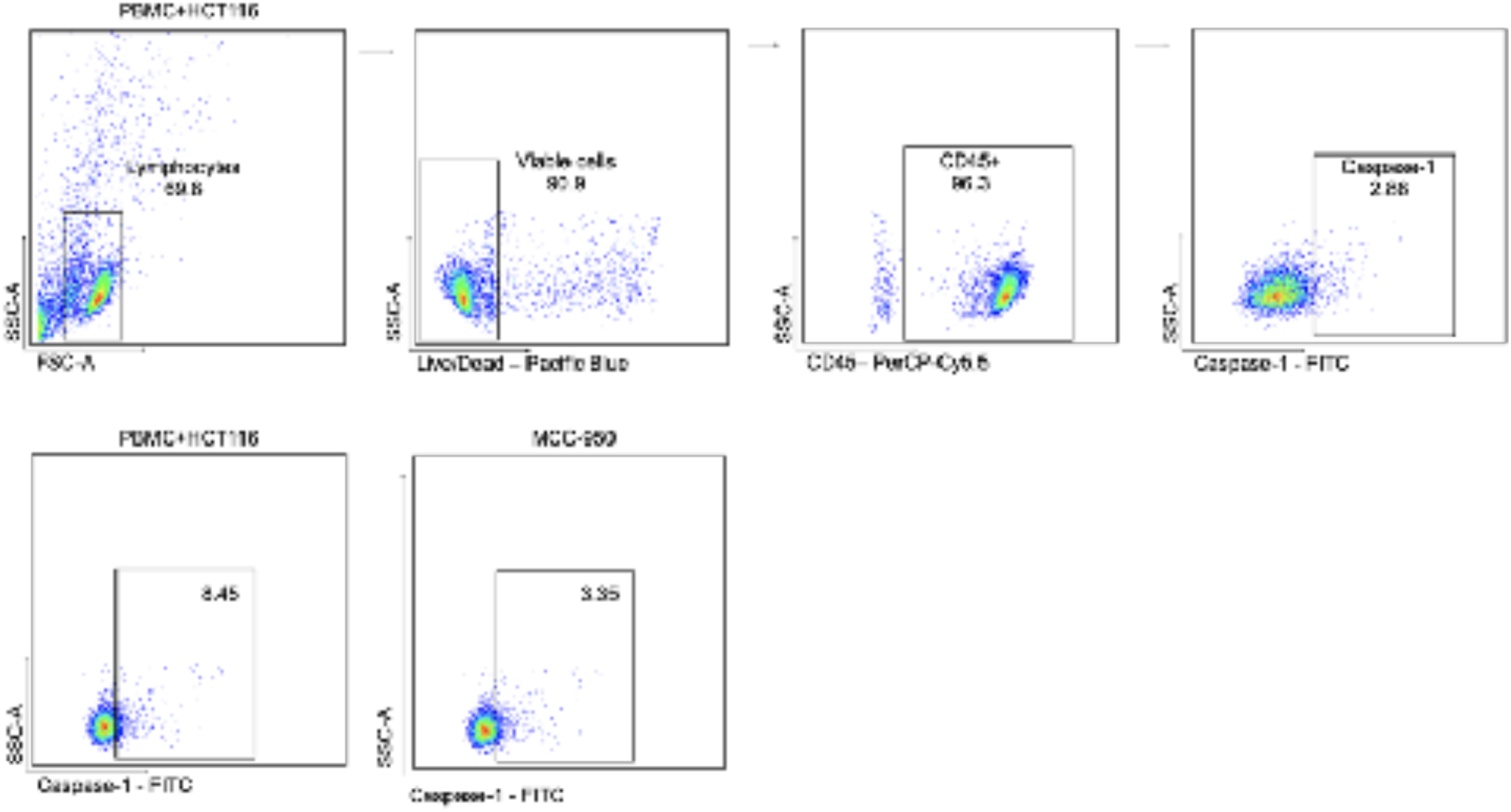
Gates strategy for flow cytometry analysis. HCT-116 spheroids (1.8 x10^5) were cultured alone or with PBMC (1.8 x10^5 cells, HCT-116/PBMC ratio: 1/1) isolated from three healthy donors for 24 hours, eventually with the pre-treatment of the NLRP3 inhibitor MCC-950 (10 µM) for 30 minutes. Cells were harvested for flow cytometry analysis. The gate strategy is reported starting from the detection of alive CD45+ cells. Within this population, the CD45+ cells positive for activated caspase-1 were selected for percentage analysis. The same gate strategy was used for the experiment with the pre-treatment with MCC-950.

**Supplementary File 4.**
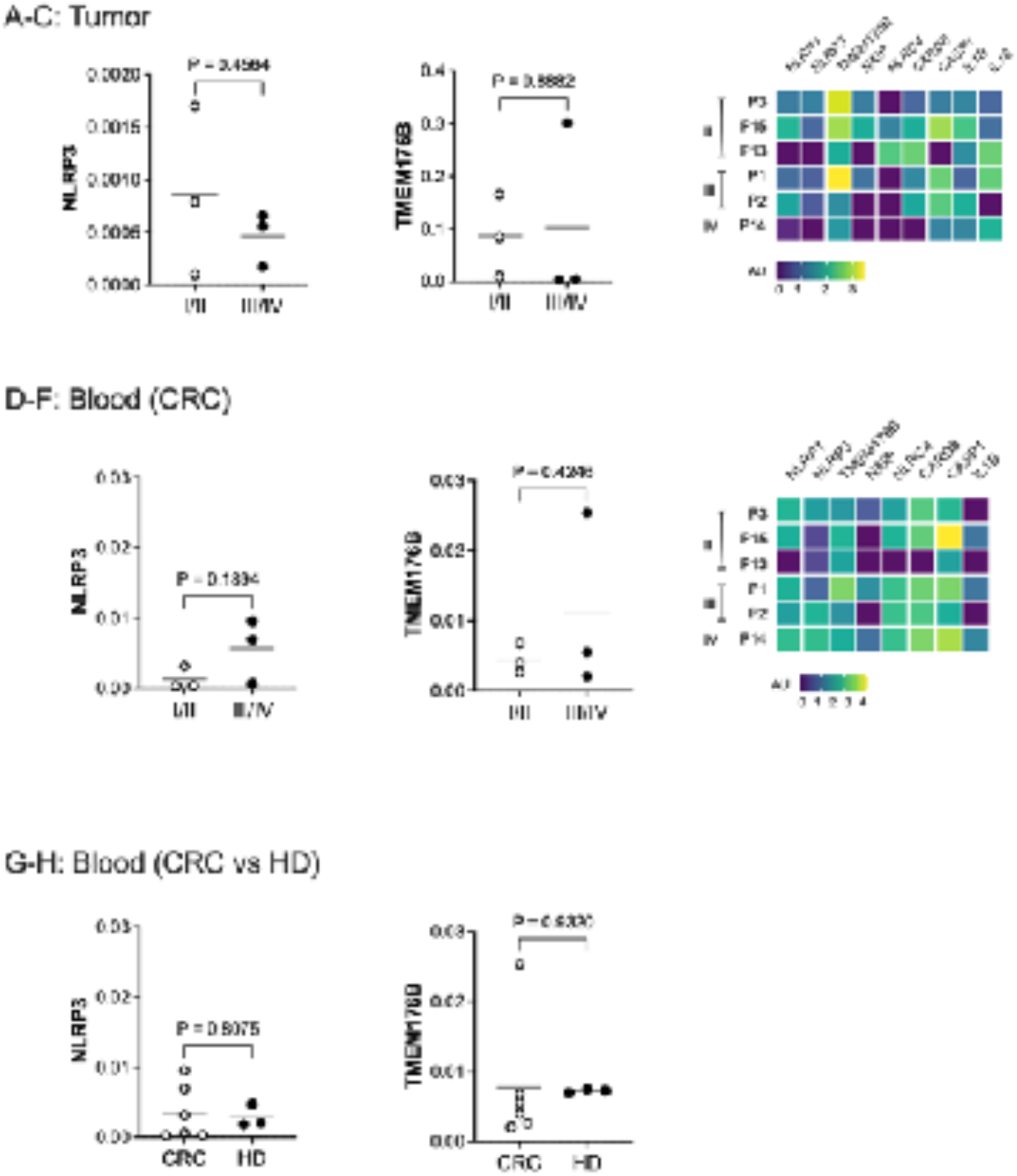
Relative expression analysis in biopsies and peripheral blood of CRC patients. Fresh biopsies and peripheral blood were collected from six heathy donors (HD) and six patients with CRC (CRC), three with a I-II TNM stage and three with a III-IV TNM stage. Total RNA were isolated from biopsies and blood “buffy-coat” and cDNA was amplified for inflammasome genes by the use of Taqman assays and qPCR. The relative expression is calculated as 2 exp -ΔCt, where ΔCt is the expression of target gene normalized for endogenous gene GAPDH. A-B) Scatter plots of NLRP3 and TMEM176B genes expression in CRC biopsies of six patients according to the stage group (TNM-II versusTNM III/IV). Values are reported as 2exp-ΔCt. Unpaired two-tails t-test p-values are indicated. C) Heatmap graph shows RNA expression level of inflammasome genes in fresh tumor biopsies of six CRC patients, three with TNM-II, two with TNM-III and one with TNM-IV. Values are reported as log10 (2exp-ΔCt). D-E) Scatter plots of NLRP3 and TMEM176B genes expression in blood leukocytes of six patients according to the stage group (TNM-II versusTNM III/IV). Values are reported as 2exp-ΔCt. Unpaired two-tails t-test p-values are indicated. F) Heatmap graph shows RNA expression level of inflammasome genes in peripheral blood leukocytes of six CRC patients, three with TNM-II, two with TNM-III and one with TNM-IV. Values are reported as log (2exp-ΔCt). G-H) Scatter plots of NLRP3 and TMEM176B genes expression in blood leukocytes of six patients and three healthy donors. Values are reported as 2exp-ΔCt. Unpaired two-tails t-test p-values are indicated.

## Notes

### Competing Interest Statement

The authors have declared no competing interest.

### Author Declarations

The Ethics Committee of ICB/USP (CEPSH) (process number 30173919.1.0000.5467) approved the study.

## REFERENCES

1. Allen IC, TeKippe EM, Woodford RM, Uronis JM, Holl EK, Rogers AB, Herfarth HH, Jobin C, Ting JP. The NLRP3 inflammasome functions as a negative regulator of tumorigenesis during colitis-associated cancer. J Exp Med. 2010 May 10;207(5):1045–56. doi: 10.1084/jem.20100050.

2. Barnett KC, Li S, Liang K, Ting JP. A 360° view of the inflammasome: Mechanisms of activation, cell death, and diseases. Cell. 2023 May 25;186(11):2288–2312. doi: 10.1016/j.cell.2023.04.025.

3. Bauer C, Duewell P, Mayer C, Lehr HA, Fitzgerald KA, Dauer M, Tschopp J, Endres S, Latz E, Schnurr M. Colitis induced in mice with dextran sulfate sodium (DSS) is mediated by the NLRP3 inflammasome. Gut. 2010 Sep;59(9):1192–9. doi: 10.1136/gut.2009.197822.

4. Cambui RAG, do Espírito Santo GF, Fernandes FP, Leal VNC, Galera BB, Fávaro EGP, Rizzo LA, Elias RM, Pontillo A. Double-edged sword of inflammasome genetics in colorectal cancer prognosis. Clin Immunol. 2020 Apr;213:108373. doi: 10.1016/j.clim.2020.

5. Cambui RAG, Fernandes FP, Leal VNC, Reis EC, de Lima DS, do Espírito Santo GF, Elias RM, Pontillo A. The Ala134Thr variant in TMEM176B exerts a beneficial role in colorectal cancer prognosis by increasing NLRP3 inflammasome activation. J Cancer Res Clin Oncol. 2023 Jul;149(7):3729–3738. doi: 10.1007/s00432-022-04284-8.

6. Condamine T, Le Texier L, Howie D, Lavault A, Hill M, Halary F, Cobbold S, Waldmann H, Cuturi MC, Chiffoleau E. Tmem176B and Tmem176A are associated with the immature state of dendritic cells. J Leukoc Biol. 2010 Sep;88(3):507–15. doi: 10.1189/jlb.1109738.

7. Deng Z, Lu L, Li B, Shi X, Jin H, Hu W. The roles of inflammasomes in cancer. Front Immunol. 2023 Jul 11;14:1195572. doi: 10.3389/fimmu.2023.1195572.

8. Domingo E, Kelly C, Hay J, Sansom O, Maka N, Oien K, Iveson T, Saunders M, Kerr R, Tomlinson I, Edwards J, Harkin A, Nowak M, Koelzer V, Easton A, Boukovinas I, Moustou E, Messaritakis I, Chondrozoumaki M, Karagianni M, Pagès F, Arnoux F, Lautard C, Lovera Y, Boquet I, Catteau A, Galon J; TransSCOT Consortium; Souglakos I, Church DN. Prognostic and Predictive Value of Immunoscore in Stage III Colorectal Cancer: Pooled Analysis of Cases From the SCOT and IDEA-HORG Studies. J Clin Oncol. 2024 Jun 20;42(18):2207–2218. doi: 10.1200/JCO.23.01648.

9. Dupaul-Chicoine J, Arabzadeh A, Dagenais M, Douglas T, Champagne C, Morizot A, Rodrigue-Gervais IG, Breton V, Colpitts SL, Beauchemin N, Saleh M. The Nlrp3 Inflammasome Suppresses Colorectal Cancer Metastatic Growth in the Liver by Promoting Natural Killer Cell Tumoricidal Activity. Immunity. 2015 Oct 20;43(4):751–63. doi: 10.1016/j.immuni.2015.08.013.

10. Galon J, Costes A, Sanchez-Cabo F, Kirilovsky A, Mlecnik B, Lagorce-Pagès C, Tosolini M, Camus M, Berger A, Wind P, Zinzindohoué F, Bruneval P, Cugnenc PH, Trajanoski Z, Fridman WH, Pagès F. Type, density, and location of immune cells within human colorectal tumors predict clinical outcome. Science. 2006 Sep 29;313(5795):1960-4. doi: 10.1126/science.1129139.

11. Galon J, Fridman WH, Pagès F. The adaptive immunologic microenvironment in colorectal cancer: a novel perspective. Cancer Res. 2007 Mar 1;67(5):1883–6. doi: 10.1158/0008-5472.CAN-06-4806.

12. Gerlo S. Interleukin-1 as Innate Mediator of T Cell Immunity. Front Immunol. 2021 Jan 27;11:621931. doi: 10.3389/fimmu.2020.621931;

13. Mailer RK, Joly AL, Liu S, Elias S, Tegner J, Andersson J. IL-1β promotes Th17 differentiation by inducing alternative splicing of FOXP3. Sci Rep. 2015 Oct 6;5:14674. doi: 10.1038/srep14674

14. Gyorffy B: Integrated analysis of public datasets for the discovery and validation of survival-associated genes in solid tumors, The Innovation, 2024, doi: 10.1016/j.xinn.2024.100625.

15. Hill M, Russo S, Olivera D, Malcuori M, Galliussi G, Segovia M. The intracellular cation channel TMEM176B as a dual immunoregulator. Front Cell Dev Biol. 2022 Oct 20;10:1038429. doi: 10.3389/fcell.2022.1038429.

16. Hill M, Russo S, Olivera D, Malcuori M, Galliussi G, Segovia M. The intracellular cation channel TMEM176B as a dual immunoregulator. Front Cell Dev Biol. 2022 Oct 20;10:1038429. doi: 10.3389/fcell.2022.1038429.

17. Huber S, Gagliani N, Zenewicz LA, Huber FJ, Bosurgi L, Hu B, Hedl M, Zhang W, O’Connor W Jr, Murphy AJ, Valenzuela DM, Yancopoulos GD, Booth CJ, Cho JH, Ouyang W, Abraham C, Flavell RA. IL-22BP is regulated by the inflammasome and modulates tumorigenesis in the intestine. Nature. 2012 Nov 8;491(7423):259–63. doi: 10.1038/nature11535.

18. Kummer JA, Broekhuizen R, Everett H, Agostini L, Kuijk L, Martinon F, van Bruggen R, Tschopp J. Inflammasome components NALP 1 and 3 show distinct but separate expression profiles in human tissues suggesting a site-specific role in the inflammatory response. J Histochem Cytochem. 2007 May;55(5):443–52. doi: 10.1369/jhc.6A7101.2006.

19. Landini G, Martinelli G, Piccinini F. Colour deconvolution: stain unmixing in histological imaging. Bioinformatics. 2021 Jun 16;37(10):1485–1487. doi: 10.1093/bioinformatics/btaa847.

20. Ruifrok AC, Johnston DA. Quantification of histochemical staining by color deconvolution. Anal Quant Cytol Histol. 2001 Aug;23(4):291–9.

21. Saponaro C, Scarpi E, Sonnessa M, Cioffi A, Buccino F, Giotta F, Pastena MI, Zito FA, Mangia A. Prognostic Value of NLRP3 Inflammasome and TLR4 Expression in Breast Cancer Patients. Front Oncol. 2021 Sep 2;11:705331. doi: 10.3389/fonc.2021.705331.

22. Schmittgen TD, Livak KJ. Analyzing real-time PCR data by the comparative C(T) method. Nat Protoc. (2008) 3:1101–8. doi: 10.1038/nprot.2008.73.

23. Segovia M, Russo S, Jeldres M, Mahmoud YD, Perez V, Duhalde M, Charnet P, Rousset M, Victoria S, Veigas F, Louvet C, Vanhove B, Floto RA, Anegon I, Cuturi MC, Girotti MR, Rabinovich GA, Hill M. Targeting TMEM176B Enhances Antitumor Immunity and Augments the Efficacy of Immune Checkpoint Blockers by Unleashing Inflammasome Activation. Cancer Cell. 2019 May 13;35(5):767–781.e6. doi: 10.1016/j.ccell.2019.04.003.

24. Segovia M, Russo S, Girotti MR, Rabinovich GA, Hill M. Role of inflammasome activation in tumor immunity triggered by immune checkpoint blockers. Clin Exp Immunol. 2020 May;200(2):155–162. doi: 10.1111/cei.13433.

25. Seo SU, Kamada N, Muñoz-Planillo R, Kim YG, Kim D, Koizumi Y, Hasegawa M, Himpsl SD, Browne HP, Lawley TD, Mobley HL, Inohara N, Núñez G. Distinct Commensals Induce Interleukin-1β via NLRP3 Inflammasome in Inflammatory Monocytes to Promote Intestinal Inflammation in Response to Injury. Immunity. 2015 Apr 21;42(4):744–55. doi: 10.1016/j.immuni.2015.03.004.

26. Shi F, Wei B, Lan T, Xiao Y, Quan X, Chen J, Zhao C, Gao J. Low NLRP3 expression predicts a better prognosis of colorectal cancer. Biosci Rep. 2021 Apr 30;41(4):BSR20210280. doi: 10.1042/BSR20210280.

27. Song-Zhao GX, Srinivasan N, Pott J, Baban D, Frankel G, Maloy KJ. Nlrp3 activation in the intestinal epithelium protects against a mucosal pathogen. Mucosal Immunol. 2014;7(4):763–774. doi:10.1038/mi.2013.94

28. Van Den Eeckhout B, Tavernier J, Gerlo S. Interleukin-1 as Innate Mediator of T Cell Immunity. Front Immunol. 2021 Jan 27;11:621931. doi: 10.3389/fimmu.2020.621931;

29. Mailer RK, Joly AL, Liu S, Elias S, Tegner J, Andersson J. IL-1β promotes Th17 differentiation by inducing alternative splicing of FOXP3. Sci Rep. 2015 Oct 6;5:14674. doi: 10.1038/srep14674.

30. Yamada SM, Pontillo A. The genetics behind inflammasome regulation. Mol Immunol. 2022 May;145:27–42. doi: 10.1016/j.molimm.2022.03.005.

31. Yang Y, Wang H, Kouadir M, Song H, Shi F. Recent advances in the mechanisms of NLRP3 inflammasome activation and its inhibitors. Cell Death Dis. 2019 Feb 12;10(2):128. doi: 10.1038/s41419-019-1413-8.).

32. Wang B, Li H, Wang X, Zhu X. The association of aberrant expression of NLRP3 and p-S6K1 in colorectal cancer. Pathol Res Pract. 2020 Jan;216(1):152737. doi: 10.1016/j.prp.2019.152737.

33. Wang P, Gu Y, Yang J, Qiu J, Xu Y, Xu Z, Gao J, Wan C. The prognostic value of NLRP1/NLRP3 and its relationship with immune infiltration in human gastric cancer. Aging (Albany NY). 2022 Dec 19;14(24):9980–10008. doi: 10.18632/aging.204438.

34. Wang Q, Hsiao JC, Yardeny N, Huang HC, O’Mara CM, Orth-He EL, Ball DP, Zhang Z, Bachovchin DA. The NLRP1 and CARD8 inflammasomes detect reductive stress. Cell Rep. 2023 Jan 31;42(1):111966. doi: 10.1016/j.celrep.2022.111966.

35. Wang Y, Gao JZ, Sakaguchi T, Maretzky T, Gurung P, Short S, Xiong Y, Kang Z. LRRK2 G2019S promotes the development of colon cancer via modulating intestinal inflammation. bioRxiv [Preprint]. 2023 Jun 30:2023.06.28.546897. doi: 10.1101/2023.06.28.546897.

36. Wu S, Zang Q, Dai B. The role of NLRP3 in the prognosis and immune infiltrates of skin cutaneous melanoma (SKCM). Transl Cancer Res. 2021 Apr;10(4):1692–1702. doi: 10.21037/tcr-20-3135.

37. Zaki MH, Vogel P, Body-Malapel M, Lamkanfi M, Kanneganti TD. IL-18 production downstream of the Nlrp3 inflammasome confers protection against colorectal tumor formation. J Immunol. 2010 Oct 15;185(8):4912–20. doi: 10.4049/jimmunol.1002046

38. Zheng L, Liu H. Prognostic association between NLRP3 inflammasome expression level and operable pancreatic adenocarcinoma. Int J Biol Markers. 2022 Sep;37(3):314–321. doi: 10.1177/03936155221096690

